# Convergent coexpression of autism associated genes suggests some novel risk genes may not be detectable in large-scale genetic studies

**DOI:** 10.1101/2022.02.28.22271620

**Authors:** Calwing Liao, Mariana Moyses-Oliveira, Celine EF De Esch, Riya Bhavsar, Xander Nuttle, Aiqun Li, Alex Yu, Nicholas D. Burt, Serkan Erdin, Jack M. Fu, Minghui Wang, Theodore Morley, Lide Han, CommonMind Consortium, Patrick A. Dion, Guy A. Rouleau, Bin Zhang, Kristen J. Brennand, Michael E. Talkowski, Douglas M. Ruderfer

## Abstract

Autism spectrum disorder (ASD) is a highly heritable neurodevelopmental disorder characterized by deficits in social interactions and communication. Protein function altering variants in many genes have been shown to contribute to ASD risk; however, understanding the biological convergence across so many genes has been difficult and genetic studies depending on presence of deleterious variation may be limited in implicating highly intolerant genes with shorter coding sequences. Here, we demonstrate that coexpression patterns from human post-mortem brain samples (N = 993) are significantly correlated with the transcriptional consequences of CRISPR perturbations (gene editing, interference and activation) in human neurons (N = 17). Across 71 ASD risk genes, there is significant tissue-specific transcriptional convergence that implicates synaptic pathways. Tissue specific convergence of risk genes is a generalizable phenomenon, shown additionally in schizophrenia (brain) and atrial fibrillation (heart). The degree of this convergence in ASD is significantly correlated with the level of association to ASD from sequencing studies (rho = -0.32, P = 3.03 ×10^−65^) as well as differential expression in post-mortem ASD brains (rho = -0.23, P = 2.39×10^−43^). After removing all genes statistically associated with ASD, the remaining positively convergent genes showed intolerance to functional mutations, had shorter coding lengths than the ASD genes and were enriched for genes with clinical reports of potential pathogenic contribution to ASD. These results indicate that leveraging convergent coexpression can identify potentially novel risk genes that are unlikely to be discovered by sequencing studies. Overall, this work provides a simple approach to functionally proxy CRISPR perturbation, demonstrates significant context-specific transcriptional convergence among known risk genes of multiple diseases, and proposes novel ASD risk gene candidates.

## Introduction

Autism spectrum disorder (ASD) is a highly heritable neuropsychiatric disorder with a population prevalence of ∼1%^1^. Sequencing studies have implicated dozens of genes contributing to risk of ASD based on an excess of rare deleterious variation in cases compared to controls^2–5^. These findings have highlighted biological pathways implicated in ASD, including synaptic function, chromatin and transcriptional regulation^2,3^. Many of these pathways have since been corroborated by transcriptomic studies^6,7,8^. However, our understanding of how these genes may interact and whether they converge on shared downstream pathways with potential to implicate novel risk genes remains incomplete.

Genetic perturbation studies involving induced pluripotent stem cells (iPSC) and CRISPR offer insight into context-specific cellular and transcriptomic consequences of perturbing ASD-associated genes individually, highlighting downstream genes or pathways that might be relevant for disease risk^9,10^. Loss of function (LoF) models of *CHD8* and *FOXP1*, two transcriptional regulators with strong association to ASD, have shown dysregulation of multiple other ASD genes^9,11^. Although several ASD risk genes have been shown to have a direct molecular relationships with each other (e.g., *CHD8* regulates and binds to other ASD risk genes like CTCF^9,12^), many altered genes lacked CHD8 and FOXP1 binding sites, suggesting that the effects of the perturbations can be propagated by downstream regulatory interactions^9,11^. In *Xenopus tropicalis*^13^, LoF CRISPR-mediated genome editing of ten ASD-implicated genes resulted in an increased ratio of neural progenitor cells to neurons in the telencephalon pointing to a convergent cellular outcome. Additional work using CRISPR-Cas9 to knock out ASD genes followed by single-cell RNA sequencing in the developing mouse brain identified recurrent glial and neuronal modules again pointing to cellular convergence^14^. Further, another study used a multiplexed iPSC platform and introduced frameshift mutations in 27 ASD genes. The ASD mutations were classified into two subgroups based on alterations in prefrontal cortex neurogenesis which then correlated with abnormal WNT signaling; one group inhibited and one group enhanced spontaneous cortical neurogenesis^15^, confirming the existence of convergent cellular and signaling phenotypes within this larger subset of ASD-associated genes.

Although these functional studies have shed some light on the interplay between multiple disease genes, determining convergent signatures across the large number of implicated ASD genes in human cell lines remains a technical and logistical challenge. These efforts are limited by the substantial time and cost required to generate human iPSCs, differentiate into a given cell type, edit the genes of interest, and investigate downstream consequences. Analysis of protein-protein interaction (PPI) networks and coexpression modules of ASD risk genes has shown that there is high connectivity, suggesting that there is more direct interaction than expected by chance^16–20^. Convergence in gene expression profiles has been demonstrated not only among ASD risk genes affected by rare protein truncation variants, but also for genes associated to ASD by large CNVs^21,22^ and common variation^23^. Similarly, ASD risk genes have been shown to be highly coexpressed in the developing cortex^18,24^. Prior work has layered expression data onto to the genetic findings to predict novel ASD risk genes^17^ successfully identifying many genes that are now significant in larger genetic studies. The high degree of connectivity across ASD risk genes presents an opportunity to leverage coexpression to better understand molecular convergence in ASD. We hypothesized that coexpression from a relevant tissue would provide a meaningful proxy for the transcriptional effects of CRISPR perturbation enabling an analysis of transcriptional convergence across many ASD risk genes to implicate novel shared downstream genes and pathways. Despite the incredible success in identifying genes for ASD, the power of genetic studies is dependent on observing variants frequently enough to ensure a statistically significant excess in cases. Therefore, power will be limited by shorter gene length or extreme genic intolerance. Implicating genes through transcriptional convergence presents one potential path to mitigating this limitation.

Here, we used coexpression to investigate the level of transcriptional convergence among ASD risk genes by leveraging large-scale post-mortem brain tissue datasets. We first showed that expression dysregulation due to CRISPR-mediated knockdown or activation is significantly and directionally correlated with coexpression with the perturbed gene demonstrating that coexpression captures downstream transcriptional consequences of perturbation. Across the set of high-confidence ASD risk genes from sequencing studies, we meta-analyzed their coexpression patterns to show highly significant tissue-specific transcriptional convergence. The generalizability of this finding is confirmed using risk genes from schizophrenia and atrial fibrillation. Our convergent ASD genes were significantly enriched for 1) neurodevelopmental disorder (NDD) genes even after excluding previous ASD-associated genes, 2) differential expression in post-mortem brains of ASD patients, and 3) synaptic function. Finally, we were able to implicate genes as novel ASD risk candidates, including smaller genes and those highly intolerant to LoF variation which precluded their identification from current genetic studies. Overall, in silico transcriptional convergence approaches as used here could be important time and cost effective additions to our understanding of disease biology.

## Methods

### CRISPR perturbation functional models

We collected data from 17 newly generated and previously published CRISPR experiments targeting 15 different ASD-associated genes. The transcriptomic data from all these CRISPR experiments were generated from isogenic iPSC-derived glutamatergic neurons induced by Neurogenin 2 (Ngn2) overexpression. Data of previously published cellular models generated by CRISPR perturbation were ascertained from the NCBI’s Gene Expression Omnibus (GEO) and Synapse. For the CRISPR-mediated LoF gene editing models described in Deneault *et al*., gene counts and covariate information were ascertained from GEO (Accession # GSE107878). These consisted of 10 ASD-relevant genes (*ANOS1, ASTN2, ATRX, CACNA1C, CHD8, DLGAP2, AFF2*/*FMR2, KCNQ2, SCN2A, TENM1*). Gene counts and covariate information for *MBD5* CRISPR-mediated LoF gene editing model were ascertained from GEO (Accession #: GSE144279). For the CRISPR activation (CRISPRa) targets (*TSNARE1, SNAP91, CLCN3*, and *FURIN*), gene counts and covariate information were ascertained from Synapse (syn20502314)^18^. CRISPR interference (CRISPRi) for *SCN2A* was ascertained from Synapse (syn26970716). From those datasets, only neuronal lines were included, and experiments were removed if there was any evidence of ineffective perturbation such as discordant RNA and protein levels of the perturbed gene.

Additionally, we used unpublished transcriptomic data from *SCN2A* and *KCTD13* CRISPR-mediated LoF gene editing models which leveraged a dual-gRNA strategy to promote gene deletions. *SCN2A* sgRNAs targeted intron 4 and intron 11 (NM_001040142) to generate a 7.5kb partial gene deletion, and the crRNA sequences used were: 5’-tatcgtagggggaccaacc-3’ and 5’-gcgtggatctagtgaactt-3’. *KCTD13* sgRNAs targeted 5’ and 3’ regions from *KCTD13* (NM_178863) to generate a 23.3kb full gene deletion, and the crRNA sequences used were: 5’-taaaaaggatggatgtaggc-3’ and 5’-tgcctgtgttaggaggtatc-3’.

The deletion lines were generated in the male control human iPSC line GM08330-8 using the Human Stem Cell Nucleofector Kit 1 (Lonza), transfecting 1μg CRISPR/Cas9 PX459 plasmid and 1μg of each gRNA using the Amaxa Nucleofection II device (Lonza), according to the manufacturer’s instructions. Cells were subsequently plated on Matrigel plates in mTeSR1 or Essential 8 medium supplemented with ROCK inhibitor for 24 hours. For clonal isolation of *SCN2A* models, puromycin selection was started 24 hours after transfection and resistant colonies were picked and expanded 48 hours after selection. For clonal isolation of *KCTD13* models, cells were separated by fluorescence-activated cell sorting (FACS) 48h after transfection. Genotyping of the resultant colonies for *SCN2A* and *KCTD13* was performed by Sanger sequencing of the deletion-specific region and ddPCR assays for copy number. A total of 4-6 successfully edited clones with heterozygous deletions plus 2-6 unedited (i.e. WT-Cas9 exposed) clonal colonies were expanded per target. Prior to neuronal differentiation, iPSC clones were split into multiple replicates, and each was manipulated in parallel during subsequent experiments.

For differentiation of *SCN2A* and *KCTD13* human iPSC models into glutamatergic neurons, Ngn2-neuronal induction was performed as previously described^25^. Briefly, iPSCs were seeded at a density of 10^6^ cells/mL and transduced with a lentivirus expressing TetO-Ngn2-GFP-Puro or TetO-Ngn2-Puro along with rtTA. Twenty-four hours after transduction, doxycycline was added to initiate Ngn2 expression, and then 24h later the cells were selected with puromycin. Ngn2-glutamatergic iPSC-derived neurons were cultured in neuronal maintenance medium supplemented with BDNF and GDNF growth factors for an additional 22 days. Subsequent experiments were performed with 24 day-old Ngn2-glutamatergic neurons, using 6-34 total replicates per genotype (i.e. WT, heterozygous deletion) per target gene.

### RNA sequencing of CRISPR perturbations

*SCN2A* RNAseq libraries were prepared from 200 ng of total RNA using a TruSeq stranded mRNA Sample Prep Kit (Illumina cat# RS-122-2102). Libraries were multiplexed, pooled and sequenced on multiple lanes of the Illumina NovaSeq platform, generating an average of 30.7M paired-end 150 bp-cycle reads for 30 samples (20 *SCN2A*^+/-^, 10 S*CN2A*^+/+^). RNAseq data was processed using a standard workflow, which includes quality assessment of fastq reads using FastQc (http://www.bioinformatics.babraham.ac.uk/projects/fastqc). Raw sequence reads were trimmed against Illumina adapters using Trimmomatic^26^ (v. 0.36) with parameters ILLUMINACLIP:adapter.fa:2:30:10 LEADING:3 TRAILING:3 SLIDINGWINDOW:4:15 MINLEN:75. Sequence reads were aligned to the human reference genome (GRCh37, Ensembl build 75) using STAR^27^ (v. 2.5.2.a) with parameters ‘–outSAMunmapped Within –outFilterMultimapNmax 1 – outFilterMismatchNoverLmax 0.1 –alignIntronMin 21 – alignIntronMax 0 –alignEndsType Local –quantMode GeneCounts –twopassMode Basic’. STAR aligner also generated gene level counts for all libraries relying on the human genome annotation provided for Ensembl GRCh37, build 75. Quality checking of alignments was assessed by custom scripts utilizing PicardTools (https://broadinstitute.github.io/picard/), RNASeQC^28^, RSeQC^29^ and SamTools^30^. The deletion introduced by CRISPR on *SCN2A* loci was validated generating exon-level coverage using DEXseq^31^ and visually investigating target loci using IGV^32^. *KCTD13* was included in gene edited as a potential contributor to 16p11.2 genomic disorder^33^, and these lines are described elsewhere^34^. In brief, RNAseq libraries were prepared using a TruSeq stranded mRNA library kit (Illumina) and were multiplexed, pooled and sequenced on multiple lanes of Illumina HiSeq 2500 platform, generating an average of 46.5M paired-end reads of 75bp for 11 samples (5 KCTD13^+/-^, 6 KCTD13^+/+^). The same RNAseq data processing pipeline without trimming step as above was applied to *KCTD13* RNAseq libraries. Processing of *MBD5* RNAseq libraries were previously described elsewhere^35^.

For the CRISPR-mediated LoF gene editing models described in Deneault et al., gene counts and covariate information were ascertained from NCBI’s Gene Expression Omnibus (GEO accession: GSE107878). The processing of samples followed standard RNA sequencing pipelines as previous described in Deneault et al. (2018)^10^.

### Differential expression analysis for CRISPR perturbations

Gene counts were input into DESeq2 for differential expression analysis. Genes with normalized counts < 10 in at least half of samples were excluded. Covariates including RNA integrity number (RIN), batch, and processing date were included when available. A Wald’s test was used for differential expression and Z-scores were calculated by dividing the fold change by the standard error. Each CRISPR perturbation was analyzed separately.

### Post-mortem brain cohorts

Two separate cohorts of post-mortem brain samples of the dorsolateral prefrontal cortex (DLPFC) were used. The CommonMind Consortium (CMC) included tissue samples from Mount Sinai NIH Brain Bank and Tissue Repository, The University of Pittsburgh NIH NeuroBioBank Brain and Tissue Repository, and the University of Pennsylvania Brain Bank of Psychiatric Illnesses and Alzheimer’s Disease Core Center. The DLPFC was dissected at each bank and sent to a centralized centre, the Icahn School of Medicine at Mount Sinai (ISMMS) for RNA extraction. Tissues from bipolar disorder (BD) or schizophrenia cases were included if they met the DSM-IV diagnostic criteria for schizoaffective disorder or schizophrenia, or for BD, which were determined in consensus conferences after reviewing of medical records, direct clinical assessments, and care provider interviews. Samples were excluded if donors had a history of Alzheimer’s disease, Parkinson’s disease, were on ventilators near time of death, or had acute neurological insults (anoxia, strokes and/or traumatic brain injury) before death.

The Human Brain Core Collection (HBCC) cohort consisted of DLPFC samples from the NIMH HBCC. The samples were analyzed clinically, neuropathologically, and toxicologically. The DSM-IV clinical diagnosis was determined through review of medical records by two psychiatrists and family interviews. Non-psychiatric controls did not have a history of substance use disorder or psychiatric conditions. Across all 933 samples, there were 345 females and 588 males. The self-reported ethnicities were 637 Europeans, 249 African, 33 Hispanic, 13 Asian and 1 other. Nearly fifty percent of samples were nonpsychiatric controls (N=462) and the remainder had a psychiatric diagnosis (113 BD, 350 schizophrenia, and 8 affective disorder)^36^. The research abided by ethical regulations and was approved by the Vanderbilt University Medical Center Review Board (IRB: 220287).

### RNA sequencing of post-mortem samples

Approximately 50 mg of homogenized tissue from the DLPFC was used to isolate RNA. The two cohorts were processed separately. Samples with age <18 were excluded prior to analysis. The RNAseq processing is identical to that described in Han et al. (2020)^37^ except for not using surrogate variable analysis (SVA) here to avoid removing trans-regulatory effects. Briefly, STAR was used to align RNA sequencing reads to GRCh37. FeatureCounts (v1.5.2) was used to count uniquely mapped reads that overlapped genes using the Ensembl v75 annotations. Fixed/mixed effects modelling were used for library normalization and covariate adjustments. Genes that were expressed at levels > 1 counts per million (CPM) in at least half of the samples in each study were retained for analysis. Conditional quantile normalization was done to account for variation in GC-content and gene length. A weighted-linear model using voom-limma was used to assess the sampling abundance confidence. Normalized log_2_(CPM) values were used for hierarchical clustering and principal component analysis to detect outlier samples. Samples were removed if deemed outliers using either method. For the CMC cohort, covariates were identified using a stepwise fixed/mixed effect regression model to identify covariates significantly associated with gene expression. The covariates were added if there was an association with principal components explaining greater than 1% of expression residual variance. For the HBCC cohort, model selection was determined through Bayesian information criteria (BIC). BIC were used to find fixed effect covariates that improved the model for most genes. Covariate adjustment was done using with a fixed/mixed effect linear regression variant, choosing mixed effect models when several samples were available per donor: gene expression ∼ covariates + sex + diagnosis + (1|Donor). Observation weights were calculated using voom-limma to adjust for the mean-variance relationship. The covariate-adjusted expression was generated after adding back the diagnostic component. This was done for both HBCC and CMC cohorts.

### Generating pairwise coexpression of genes in the DLPFC

For each of the two cohorts, the covariate-adjusted expression was used to calculate coexpression values across all 16,992 genes. A pairwise Pearson’s correlation was calculated for each pair of genes and the correlation coefficient was subsequently transformed into a Z-score using a Fisher transformation. The coexpression Z-scores for each pair of genes were subsequently meta-analyzed across the two cohorts using Stouffer’s weighted Z-score method.

### Assessing relationship between CRISPR perturbation and coexpression

For each experiment, we calculated Pearson correlation between the rank normalized differential expression and the perturbed gene’s coexpression profiles from the post-mortem brain tissue. ASD CRISPR perturbations were meta-analyzed using Stouffer’s weighted Z-score and compared to the meta-analyzed coexpression of the same genes. The relationship was subsequently assessed with a Pearson’s correlation.

### ASD convergent coexpression meta-analysis

We included 71 genes implicated in risk of ASD from a cross-consortia exome sequencing study that combined *de novo* and inherited single-nucleotide variant (SNV), indel, and CNV analyses (FDR < 0.001, approximating exome-wide Bonferroni correction)^5^. After filtering out genes due to low expression level, we performed a meta-analysis of 71 genes. Coexpression Z-scores were meta-analyzed using Stouffer’s weighted Z-score method to generate convergent effect sizes. To assess whether convergence of ASD risk genes is tissue specific, we used Genotype Tissue Expression Consortium (GTEx) RNAseq counts of the frontal cortex (BA9) (N=209), left ventricle (N=432), skeletal muscle (N=803) and liver (N=226) as negative controls. We selected these as they are not derived from the ectoderm or previously implicated in ASD. Convergence was calculated using the same methods as described previously. To assess the null distribution for ASD convergence, we conducted 10,000,000 permutations where for each permutation a meta-analysis was performed using 71 randomly-selected genes, excluding the 71 ASD genes. The empirical p-values were calculated, as shown here: (# of absolute convergent Z-scores greater than or equal to the absolute convergent ASD Z-score + 1) / (Total # of permutations + 1). A Bonferroni-correction was then applied to the empirical p-values and a threshold of P < 0.01 after correction was used to increase stringency.

To account for the potential effects of confounding from LoF observed/expected upper bound fraction (LOEUF), the permutation was repeated by matching on LOEUF scores within -/+ 0.05 of each gene. Genes that were Bonferroni-significant and large effect (absolute Z-score > 2) were input into ToppGene for pathway enrichment, with the background gene set being all unique genes with coexpression values. Afterwards, we sought to determine whether the convergent coexpression relates to differentially expressed ASD genes. Summary statistics from the ASD vs control post-mortem brain PsychENCODE dataset were ascertained and correlated against the convergent coexpression.

### Dissecting relationship between convergence, intolerance and ASD association

Since ASD risk genes are strongly intolerant and intolerant genes are more coexpressed with each other, we wanted to assess whether among the significant convergent genes were also tolerant genes associated with ASD. To define intolerance, we divided the genes into two sets, tolerant (LOEUF >0.35) and intolerant (LOEUF <0.35). The correlation was assessed between transcriptional convergence and the -log_10_(p) significance of the exome data. Next, we sought to identify whether transcriptional convergence can identify novel ASD genes that have not been previously implicated due to limitations of genetic studies. We defined ASD association as having a total Bayes factor (BF) > 2 for the exome data which is includes the presence of even a weak association. A BF of greater than 3 is typically used to represent meaningful significance. We assessed the correlation between LOEUF and transcriptional convergence for both associated and non-associated ASD genes.

### Classification of 71 ASD associated genes

We categorized 71 ASD associated genes into three main functional groups including chromatin, transcription and synaptic by their association with chromatin function related GO terms (“GOMF_CHROMATIN_BINDING”, “GOMF_CHROMATIN_DNA_BINDING”, “GOBP_REGULATION_OF_CHROMATIN_ASSEMBLY_OR_DISASSEMBLY”,”GOBP_CHROMATIN_ ORGANIZATION”,”GOBP_CHROMATIN_REMODELING”,”GOBP_REGULATION_OF_CHROMATIN_ BINDING”,”GOBP_CHROMATIN_MEDIATED_MAINTENANCE_OF_TRANSCRIPTION”,”GOBP_CHR OMATIN_MAINTENANCE”,”GOBP_REGULATION_OF_CHROMATIN_ORGANIZATION”) and transcription related GO terms (“GOBP_MRNA_TRANSCRIPTION”, “GOBP_REGULATION_OF_TRANSCRIPTION_REGULATORY_REGION_DNA_BINDING”, “GOBP_MRNA_TRANSCRIPTION_BY_RNA_POLYMERASE_II”,”GOBP_CHROMATIN_ORGANIZATI ON_INVOLVED_IN_REGULATION_OF_TRANSCRIPTION”,”GOMF_RNA_POLYMERASE_II_TRANS CRIPTION_FACTOR_BINDING”,”GOMF_TRANSCRIPTION_COREGULATOR_BINDING”,”GOMF_DN A_BINDING_TRANSCRIPTION_FACTOR_ACTIVITY”,”GOMF_TRANSCRIPTION_FACTOR_BINDIN G”,”GOMF_TRANSCRIPTION_REGULATOR_ACTIVITY”) and synaptic function based on manual curation reported in SynGO v1.1 database^38^. The above listed GO terms and their associated genes were retrieved from MSigDB (v7.4) database. Genes that were not identified in any of these categories were manually further classified based on https://www.genecards.org/.

## Results

### Characterization of CRISPR-mediated perturbations

RNA-sequencing data from 17 CRISPR perturbation experiments in neurons comprising 15 unique genes were included from three independent sources (see Methods). In total, the CRISPR experiments included: twelve CRISPR-mediated heterozygous or homozygous LoF mutational models (*AFF2, ANOS1, ASTN2, ATRX, CACNA1C, CHD8, KCTD13, KCNQ2, MBD5, SCN2A [2x]*, and *TENM1)*^10,35^, four CRISPR activation models (*FURIN, SNAP91, TSNARE1*, and *CLCN3)*^39^, and one CRISPR interference model (*SCN2A)*. We tested for differential expression between the CRISPR edited cell lines and the unedited cell lines for each gene passing quality control including observing the expected effect in the perturbed gene (see Methods). Each experiment was analyzed separately, and the differential expression effect sizes were converted to Z-scores representing “CRISPR perturbation.” The experiments had variable impact on global expression patterns, with the total number of significantly differentially expressed genes identified ranging from 164 to 4,857 and lambda inflation factors ranging from 0.29 – 4.13 (Table 1).

**Table 1.** CRISPR modelling experimental design and numbers of differentially expressed genes per experiment. *Dataset described in Deneault *et al*.^10^ Hem: Hemizygous, Hom: Homozygous, Het: Heterozygous.

| Gene | Type of CRISPR perturbation | Number of Edited Lines | Number of Unedited Lines | Number of significant negative genes (P<0.05) | Number of significant positive genes (P<0.05) | Number of significant genes (P<0.05) | Lambda |
| --- | --- | --- | --- | --- | --- | --- | --- |
| *AFF2* * | Hem gene edit | 5 | 4 | 151 | 146 | 297 | 0.48 |
| *ANOS1* * | Hem gene edit | 4 | 4 | 122 | 42 | 164 | 0.29 |
| *ASTN2* * | Hom gene edit | 3 | 4 | 455 | 173 | 628 | 0.75 |
| *ATRX* * | Hem gene edit | 4 | 4 | 991 | 966 | 1957 | 1.39 |
| *CACNA1C* * | Hom gene edit | 3 | 4 | 166 | 132 | 298 | 0.57 |
| *CHD8* * | Het gene edit | 4 | 4 | 112 | 107 | 249 | 0.5 |
| *KCNQ2* * | Hom gene edit | 4 | 4 | 381 | 329 | 710 | 1.29 |
| *KCTD13* | Het gene edit | 6 | 5 | 2167 | 2690 | 4857 | 4.13 |
| *MBD5* | Het gene edit | 6 | 3 | 81 | 177 | 258 | 0.72 |
| *SCN2A* | Het gene edit | 20 | 10 | 573 | 1137 | 1710 | 1.84 |
| *SCN2A* * | Hom gene edit | 4 | 4 | 143 | 80 | 223 | 0.53 |
| *SCN2A* | CRISPRi | 2 | 2 | 2062 | 2525 | 4587 | 2.13 |
| *TENM1* * | Hem gene edit | 5 | 4 | 1114 | 1280 | 2394 | 2.33 |
| *CLCN3* | CRISPRa | 6 | 6 | 1049 | 605 | 1654 | 1.31 |
| *FURIN* | CRISPRa | 2 | 2 | 399 | 268 | 667 | 0.54 |
| *SNAP91* | CRISPRa | 2 | 2 | 434 | 764 | 1198 | 1.11 |
| *TSNARE1* | CRISPRa | 2 | 2 | 626 | 825 | 1451 | 1.07 |

One gene (*SCN2A*) was perturbed in three independent experiments allowing us to quantify the variability of global gene expression changes due to CRISPR perturbation and genetic background. We found significant but modest Pearson correlations of differential expression across experiments that varied from 0.202 between the two CRISPR-mediated gene editing experiments to 0.343 and 0.335 between the CRISPRi experiment and the two gene edit experiments (Supplementary Figure 1).

### Gene coexpression correlates with downstream transcriptional consequences of CRISPR perturbation

We hypothesized that gene coexpression would proxy differential expression driven by CRISPR perturbations in the experiments described above. We leveraged 993 post-mortem brain samples from the dorsolateral prefrontal cortex (DLPFC) and calculated pairwise gene coexpression using Pearson correlation (see Methods). We identified consistent and significant negative correlation between differential expression due to CRISPR-mediated gene knockdown and the corresponding perturbed gene’s normalized coexpression profile ranging from -0.089 to -0.45 (Figure 1A). The exception was the *ANOS1* LoF model which had a nominally significant positive correlation (R=0.018, P=0.046). Overall, genes that are more significantly downregulated by a specific CRISPR perturbation are more highly and positively coexpressed with the perturbed gene. This holds true when CRISPR targets are downregulated via LoF mutation or CRISPRi as well as when they are upregulated using CRISPRa (Supplementary Figure 2).

**Figure 1.**
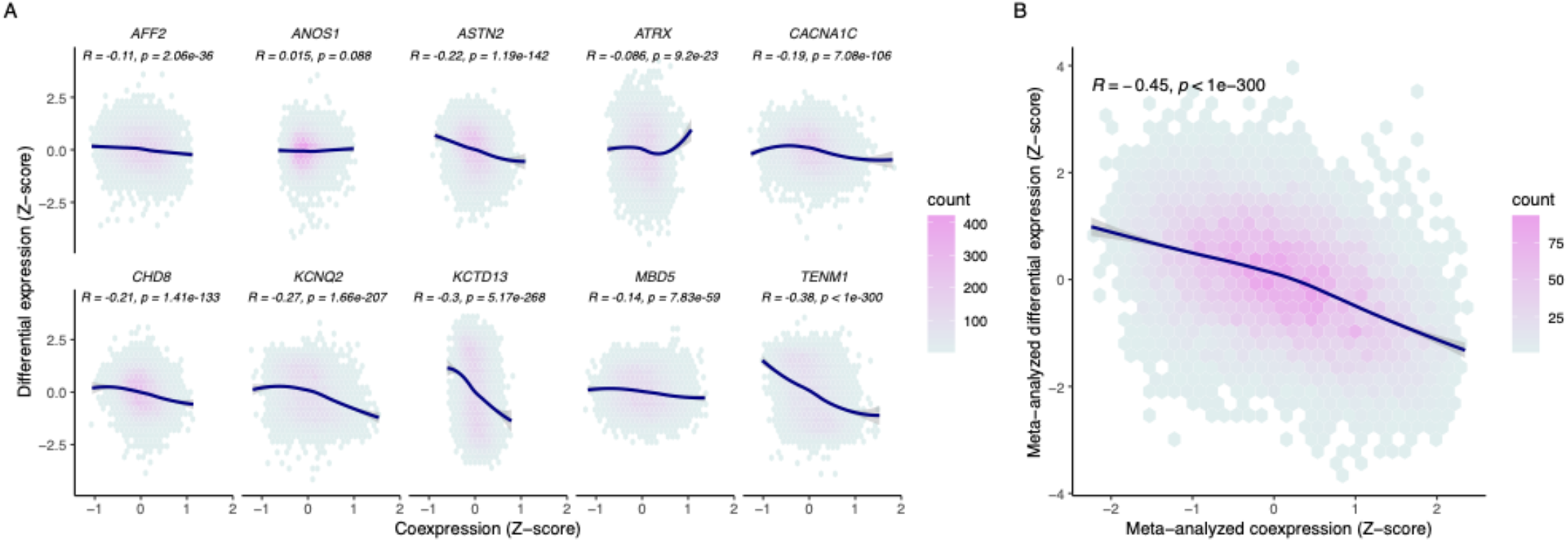
Coexpression proxies CRISPR-induced differential expression. (A) Correlation between differential expression profiles for ten CRISPR knockdown models (y-axis) with respective coexpression profiles (x-axis) for each gene (points). (B) Meta-analysis across 10 CRISPR perturbations with respective meta-analyzed coexpression. Coexpression is represented as a Fisher transformed Pearson’s correlation Z-score. A Pearson correlation was done to assess the correlation between coexpression and differential expression. The curve for each panel was fitted with a locally weighted smoothing (LOESS) regression.

With a variably sized but consistently significant negative relationship seen across single gene CRISPR perturbation and coexpression, we next asked whether meta-analyzing differential expression from multiple CRISPR perturbations could also be proxied by a meta-analysis of the coexpression profiles of the perturbed genes. In other words, we wanted to assess whether genes that were consistently differentially expressed in the same direction across multiple gene perturbations (convergent genes) could be inferred from a similar convergence of coexpression of the respective genes from post-mortem brain tissue. When we separately meta-analyzed all CRISPR-mediated gene edits and corresponding coexpression profiles for the perturbed genes, we found that convergent CRISPR perturbation was significantly negatively correlated with convergent coexpression (Pearson’s R = -0.44, P < 1×10^−300^, Figure 1B).

Given the significant correlation between CRISPR perturbation and coexpression, we next assessed how that compared to the correlation of differential expression from different CRISPR perturbations of the same gene (*SCN2A*) across three different experiments. Significant Pearson correlation was observed between *SCN2A* coexpression and differential expression in each CRISPR experiment of *SCN2A* ranging from -0.25 – -0.43 (Figure 2). The correlation statistics across CRISPR *SCN2A* experiments (0.2 – 0.34) were similar to those seen when compared to coexpression suggesting that coexpression provided a similar proxy to transcriptional dysregulation as an independent CRISPR experiment. After meta-analyzing the differential expression profiles from the three *SCN2A* CRISPR experiments, the correlation with coexpression was stronger than each individual experiment alone (Pearson’s R = -0.45, P < 1×10^−300^).

**Figure 2.**
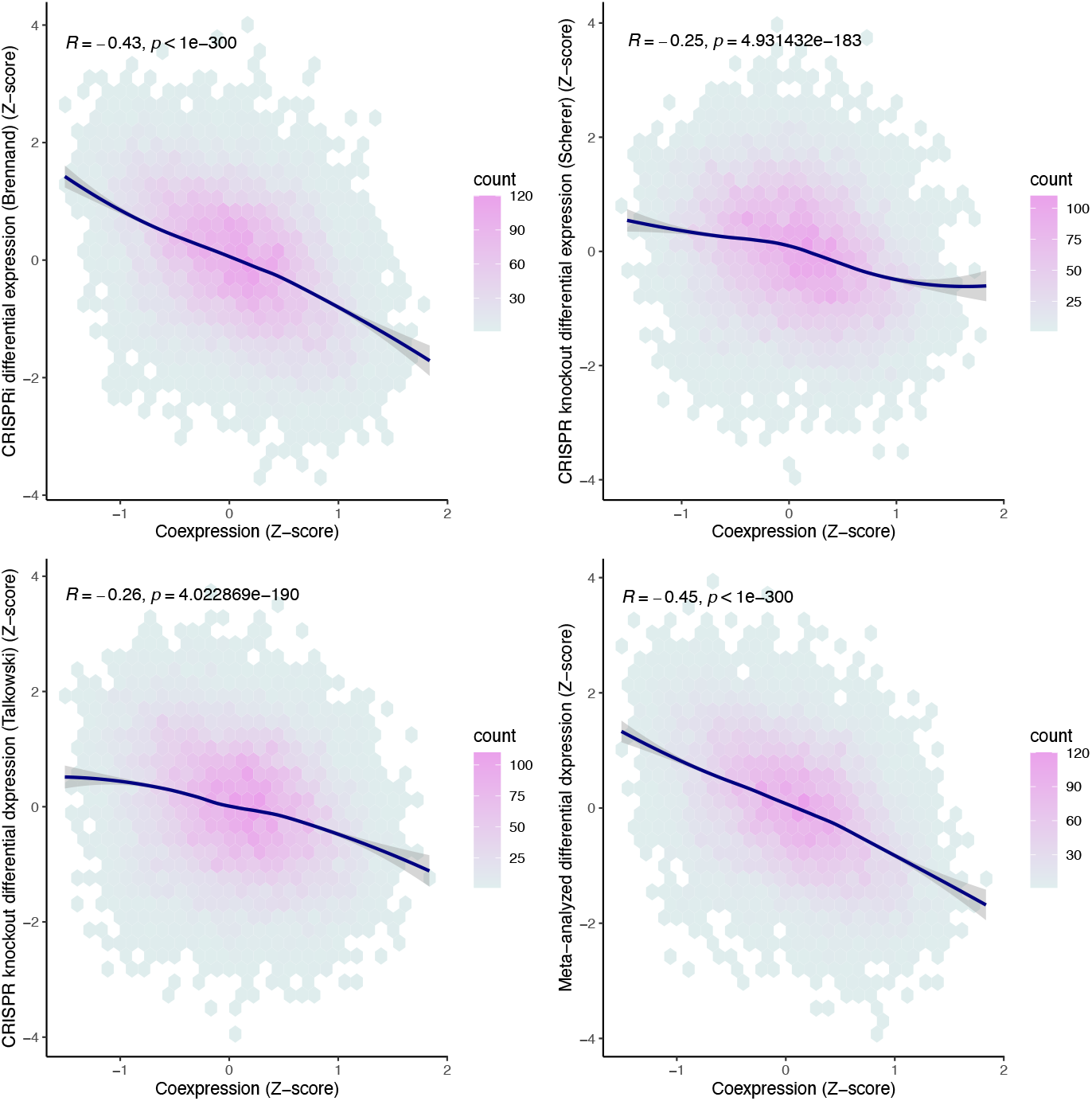
Coexpression consistently proxies differential expression for *SCN2A* across three independent perturbations. Coexpression represented as a Fisher transformed Pearson’s correlation Z-score (x-axis). A Pearson correlation was calculated to assess the correlation between coexpression and differential expression (y-axis) for each gene (points). The curve for each panel was fitted with a locally weighted smoothing (LOESS) regression.

### In silico functional convergence of ASD risk genes

We next sought to leverage coexpression to test convergence of the 71 risk genes implicated from the most recent ASD exome sequencing study^5^. After calculating meta-analysis effect sizes of Fisher’s transformed Pearson’s correlations, we assessed significance by running 10,000,000 permutations where the coexpression of 71 randomly selected genes (among those with coexpression) were meta-analyzed (see Methods). Significant overall convergence was identified by comparing the variance of the distribution of convergent coexpression of the ASD genes to the variance of the permuted random genes (ASD variance = 1.70, mean permuted null variance = 0.131, P = 9.9×10^−7^). A total of 3,553 genes were found to be significantly convergently coexpressed at a Bonferroni-adjusted empirical p-value less than 0.01. Nearly 60% of those (n=2,157) were positively convergent suggesting that their expression was consistently in the same direction as the ASD genes. Comparison to convergence defined using the rank-based Spearman’s correlation yielded consistent results and near total correlation (Spearman’s Rank Correlation, rho = 0.995, P < 1×10^−300^).

The analysis that identified the 71 ASD genes directly incorporated constraint against LoF variation (LOEUF^40^) into estimates of prior relative risk, which could inflate coexpression convergence among intolerant genes. To account for this potential confounding, we reran the permutation procedure randomly selecting 70 genes with matching LOEUF scores to the ASD genes. The p-values between intolerance-matched permutation and not were strongly correlated (Spearman’s Rank Correlation, rho = 0.93, P < 1×10^−300^), suggesting that the models of ASD risk gene discovery did not significantly inflate discovery of convergent genes (Supplementary Figure 3). Further, an analysis based on a Fisher’s combined test using only *de novo* and missense variants without incorporating intolerance identified 49 ASD genes (FDR < 0.001) that showed no significant difference in intolerance scores compared to the 71 ASD genes (Two-sided Wilcoxon Test, W = 1825.5, P = 0.648). Given that intolerance did not seem to confound our analyses, we meta-analyzed the initial 71 genes for downstream analyses.

Convergent genes were overrepresented in gene sets related to neuronal and synaptic function with the most significant pathways across categories including the synapse (P = 1.79×10^−21^), synaptic signaling (P = 6.23×10^−19^), neuronal system (P = 1.74×10^−13^) and abnormal CNS synaptic transmission (P = 3.03×10^−13^) (Supplementary Table 1). Convergent genes also represented 127 of the 410 genes implicated in neurodevelopmental disorders from DisGeNet^41^ (P = 2.96×10^−7^) and 71 of the 193 genes in the cation channel complex (P = 1.45×10^−7^). Next, we partitioned the significantly convergent genes by direction of effect and found that positively convergent genes were largely enriched in synaptic pathways (Supplementary Table 2) whereas negatively convergent genes had much less clear pattern of enrichment across pathways (Supplementary Table 3).

### Convergence captures ASD signal from exome and post-mortem brain studies

Given the strong enrichment of synaptic functions observed across convergent genes and the neurodevelopmental deficits observed in ASD, we reasoned that convergent genes contributing to ASD etiology would display disease-relevant tissue specificity. We tested this hypothesis using GTEx^42^ data and found no meaningful convergence in liver, left ventricle, or muscle coexpression, however, we saw convergence in frontal cortex tissue (Figure 3A). The mean absolute convergence Z-score for the liver, heart and muscle were 0.318, 0.655, and 0.591 respectively, while the mean absolute Z-score from the DLPFC was 1.05 and was significantly different than the non-brain tissues (Two-sided Wilcoxon Test, P < 2.2×10^−16^), suggesting tissue specificity of ASD convergence. The mean absolute convergence Z-score for the frontal cortex was 1.10, which was similar to the DLPFC. To determine if the enrichment of convergent genes associated with synaptic biology was driven by ASD risk genes associated with synaptic functions, we assigned 60 of the 71 ASD genes to relevant pathway terms associated with synaptic, transcription factor, and chromatin biology. The convergence permutations were rerun for each set separately and we found that convergence was strongly driven by synaptic genes, whereas the chromatin genes and transcription factors displayed progressively weaker convergence (Synaptic variance: 2.57, chromatin variance = 0.39, transcription variance = 0.18) (Supplementary Figure 4).

**Figure 3.**
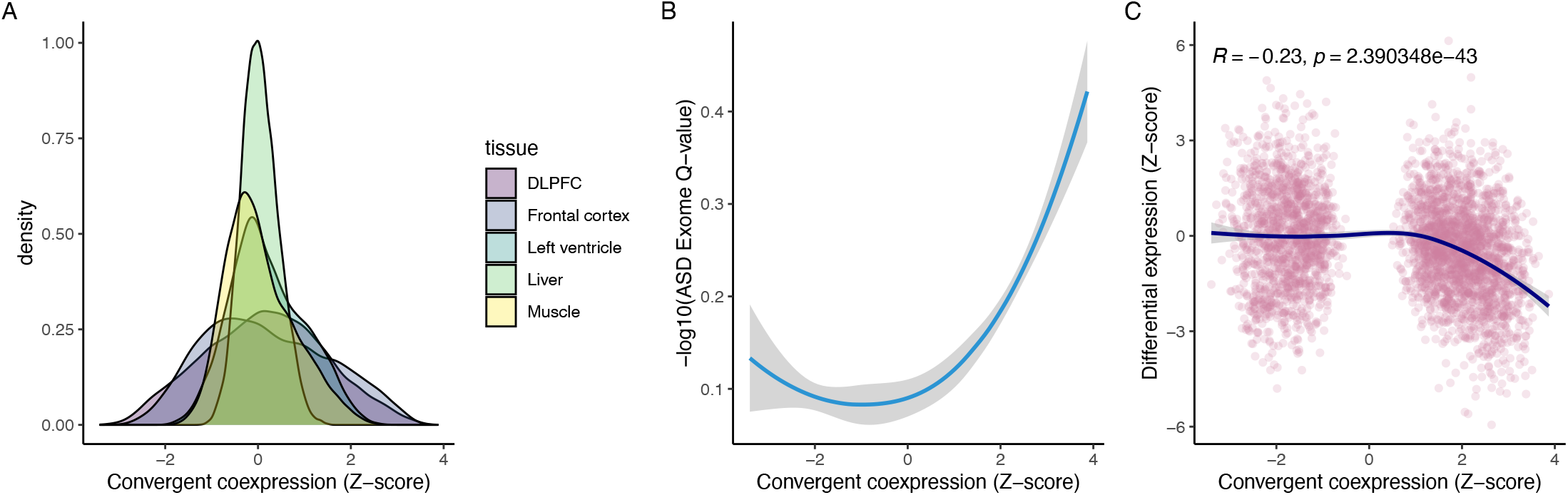
Convergence is tissue specific and associated with ASD. (A) Distributions of transcriptional convergence of 71 high-confidence ASD genes in different tissues. (B) Smoothed relationship with confidence interval between ASD exome significance and significantly convergent coexpression effect sizes (P_bonf_ < 0.01). (C) Correlation between convergently coexpressed genes (P_bonf_ < 0.01) and differential expression of ASD post-mortem DLPFC compared to controls. The curve was fitted with a locally weighted smoothing (LOESS) regression.

To assess the generalizability of this approach, we quantified convergence amongst 10 schizophrenia (SCZ) risk genes identified by SCHEMA^43^. We found significant convergence in the DLPFC (absolute mean = 0.74, variance = 0.81), but not in the unrelated tissues such as the left ventricle, muscle and liver. Next, we asked whether convergent genes overlapped between SCZ and ASD, given known overlap in disease biology. Among the 135 significant SCZ convergent genes, 124 were also significant ASD convergent genes (Fisher’s Exact Test, OR: 60.80 [29.9 – 144.0], P = 2.43 × 10^−74^). Then, we selected an unrelated non-brain phenotype in atrial fibrillation and found no convergence in the brain tissues, but 3040 significant convergent genes (P_bonf_ < 0.01) in the left ventricle (Supplementary Figure 5).

Finally, we asked whether the ASD convergent genes could inform on the biology of ASD. Excluding the 71 ASD genes used to calculate convergence, there was a significant correlation between convergence and evidence for ASD risk based on the significance of each of the remaining genes to ASD (Spearman’s Rank Correlation, rho = -0.316, P = 3.03×10^−65^) (Figure 3B). There was a significant enrichment of ASD genes implicated through rare variant analyses (q-value <0.05) among the positively convergent genes (Z > 2) (OR:

4.63 [2.97 – 7.10], P = 1.02×10^−10^). This effect size was larger when only including Bonferroni-significant (P < 0.01) positively convergent genes (OR: 7.41 [3.10 – 20.40], P = 1.85×10^−7^). That is, genes most positively convergent were also most likely to be associated to ASD through rare variant analyses. This finding was also seen among SCZ convergence and the SCHEMA association results (Spearman’s Rank Correlation, rho = -0.276, P = 0.005).

ASD risk genes are biologically more likely to be intolerant. Convergent genes also are significantly more likely to be intolerant (mean LOEUF for convergent genes = 0.73, mean LOEUF for other genes 0.83, Wilcoxon p-value = 2.85 × 10^−31^). However, after splitting genes based on intolerance, we found a significant correlation between convergence and ASD association for both tolerant (LOEUF > 0.35, Spearman’s Rank Correlation, rho=0.18, P=1.91×10^−15^) and intolerant genes (LOEUF < 0.35, Spearman Rank Correlation, rho=0.14, P=8.57×10^−5^) (Supplementary Figure 6). A significant relationship also existed between convergence and differential expression of ASD post-mortem brain tissue compared to controls with the positive convergence being correlated with downregulation in ASD (Spearman’s Rank Correlation = -0.23, P = 2.39×10^−43^, Figure 3C).

### Potential to identify novel ASD genes missing from current genetic analyses

Genes with shorter coding sequences will have reduced power in genetic analyses that require multiple deleterious variants among cases to quantify risk. Similar issues could exist for genes where deleterious variation is inviable but less deleterious modulation could contribute to ASD risk. Given the significant relationship between convergence and ASD association, we asked whether our set of convergent genes included potential ASD risk genes with properties that current genetic studies might be underpowered to identify. Among the most significantly associated 71 ASD risk genes, there was a highly significant skew towards longer coding sequence (median = 3,642bp) compared to 1,293bp among all other genes. After splitting our positive convergent genes into those with even a weak association to ASD (BF > 2) or not, we identified significantly increased median coding sequence lengths in the ASD associated genes compared to the rest (median ASD-associated coding sequence length = 2,723 bp; median non-ASD -associated coding sequence length = 1,809 bp, P = 5.76×10^−25^ Wilcoxon Signed Rank Test, Figure 4A). The most positively convergent genes not associated to ASD had similar intolerance scores to those that were linked to ASD (Figure 4B). Consistent results were identified among convergent SCZ genes (Supplementary Figure X). We show that positive convergence is more significantly correlated with the ASD effect size driven by missense than protein-truncating variation (Spearman’s Rank Correlation, missense type A, missense variants with missense badness, Polyphen-2, and constraint [MPC]^44^ scores between 1 and 2 : rho = 0.117, P = 5.36×10^−05^; missense type B, missense variants with MPC scores greater than or equal to 2: rho = 0.103, P = 3.96×10^−04^; protein-truncating: rho = 0.04, P = 0.039).

**Figure 4.**
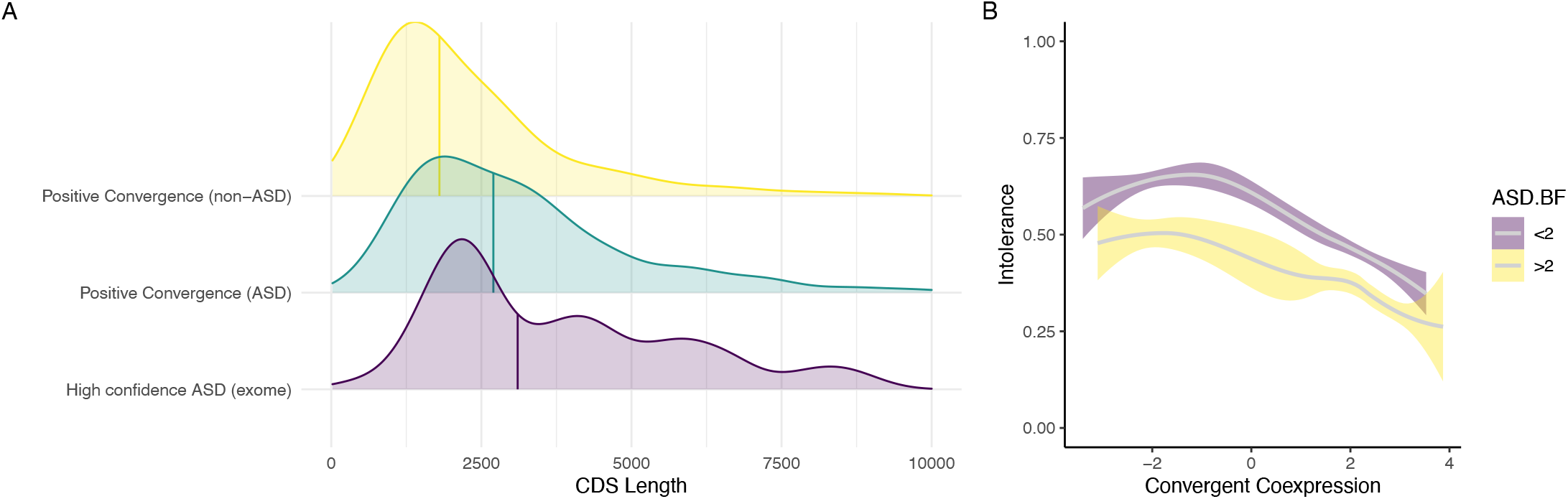
Convergence captures novel intolerant and small genes implicated in ASD risk. (A) Distributions and medians (colored vertical lines) of coding sequence lengths for significant positively convergent genes stratified by their association with ASD. High confidence ASD exome genes (purple) consist of the 71 genes with a q-value < 0.001. A Bayes factor (BF) > 2 in the ASD exome data is considered ASD-associated. The positive convergent genes are split by those with BF > 2 (green) and those with BF < 2 (yellow). (B) Relationship between convergent coexpression and intolerance (LOEUF scores) stratified by association with ASD amongst significantly convergent genes. Purple represents ASD non-associated genes (BF < 2) and yellow indicates ASD-associated genes (BF > 2). Significant convergence defined as P_bonf_ < 0.01.

Finally, we sought to assess whether convergence may increase power to detect genes that have some level of evidence linking them to ASD risk but have not been implicated in exome-consortia analyses. To address this question, we leveraged the manually curated database of genes implicated in ASD susceptibility by the Simons Foundation Autism Research Initiative (SFARI, https://gene.sfari.org/). Genes in this database come from large-scale sequencing studies but also from functional studies, clinical reports and genetic studies of common variation. After excluding all genes associated with ASD using the same criteria as before (BF > 2), there was a significant enrichment of the remaining positively convergent genes among this broader ASD geneset (Fisher’s Exact Test, OR: 3.00 [2.07 – 4.37], P = 1.00×10^−09^) (Supplementary Table 4). This finding suggests that convergence may provide a useful supplement to sequencing studies in searching for additional ASD risk genes.

## Discussion

Understanding the context-specific functional consequences of perturbing genes will be important in elucidating the molecular underpinnings of disease. Since *in vitro* experiments remain costly and challenging to scale, *in silico* approaches provide immediate opportunities to improve understanding. Here, we show that coexpression can proxy the regulatory consequences of CRISPR perturbation across shared contexts with similar correlation to replicate CRISPR experiments involving the same gene. Post-mortem brain coexpression meta-analyzed across 71 genes implicated in ASD was then used to demonstrate highly significant regulatory convergence among synaptic genes. Finally, these convergent genes were found to be enriched for genes with support linking them to ASD but without definitive statistical evidence from large-scale genetic studies, which are underpowered to evaluate genes with short coding sequence length and high intolerance.

Our hypothesis was that gene-gene coexpression would capture some proportion of the downstream transcriptional consequences of perturbing a gene through CRISPR in a shared context, providing an opportunity to “proxy” these effects *in silico* and assess functional effects shared across many disease genes (“convergence”). Regardless of whether a CRISPR target gene was upregulated or downregulated, genes that were positively coexpressed with the target gene experienced transcriptional dysregulation in the same direction as the target. For CRISPRi gene knockdown experiments, genes positively coexpressed with the perturbed gene showed decreased expression, and for CRISPRa, such genes showed increased expression. Importantly, we show that the correlation of coexpression and CRISPR perturbation is similar to the correlation of different CRISPR experiments modulating the same gene. Furthermore, the meta-analysis incorporating multiple CRISPR experiments is even more highly correlated with coexpression, highlighting the variability in perturbation across CRISPR experiments and potential benefits of repeated experiments with different variants and backgrounds. Additionally, we found that the convergent differential expression from multiple CRISPR perturbations could be inferred by convergent coexpression of the same perturbed genes. These results suggest that coexpression could be used to assess transcriptional convergence of disease-relevant genes.

We explored this possibility by assessing the transcriptional convergence of 71 ASD risk genes and found that there was highly significant convergence. The degree of convergence was context-specific, absent in coexpression data from liver, left ventricle, or muscle but with a strong signature in both brain tissues. We found context specific convergence existed in other brain and non-brain diseases such as SCZ and atrial fibrillation demonstrating generalizability of the approach. Significant convergence was also correlated with evidence of association to ASD from sequencing studies and with differential expression between ASD cases and controls in post-mortem brain tissue. Moreover, there are multiple pathways relevant to neurodevelopmental disorders significantly enriched for the convergent genes, including pathways involving the synapse and ion channels^45–47^, reinforcing the link to ASD. Intriguingly, the relationship between convergence and ASD is predominantly a product of positive convergence, or genes positively coexpressed with many of the ASD risk genes. In general, there are more strongly positively convergent genes than negative. However, despite there being many significant negative convergent genes, these genes as a class lack existing evidence for ASD association, and compared to the positive convergent genes, they do not show enrichment within previously implicated ASD pathways. These results could point to a bias of coexpression, reflect ascertainment limitations arising from genes harboring of *de novo* LoF variants, or signal a biological phenomenon where only downstream functional effects in the same direction as dysregulation of risk genes contributes to risk.

Large-scale genetic studies have contributed dramatically to our current knowledge regarding the biological basis of ASD and spearheaded the identification of the risk genes used here to quantify convergence. These studies depend on observing a statistically significant excess of deleterious variation among cases. Factors that reduce the likelihood of finding variants such as short coding sequence or mutational inviability diminish the power of these studies, potentially precluding genuine ASD risk genes from discovery via this approach. We were interested in whether our convergence metric could identify such putative “hidden” ASD risk genes. We show that our convergent genes are substantially shorter but similarly intolerant compared with genes previously associated with ASD and SCZ. Finally, we show that convergent genes not associated with ASD from sequencing studies are still enriched for genes implicated in ASD from clinical diagnostic studies, functional studies or analyses of common variation nominating our convergent genes described here as potential novel ASD risk genes. For example, *LMTK2*, was the most convergently coexpressed gene with no association to ASD. It is highly intolerant to LoF mutation (LOEUF = 0.24) and highly expressed in brain tissue^40,42^. Interestingly, disruption of this gene contributes to infertility phenotypes in male mice^48^. The gene has also been linked to Alzheimer’s disease and has been suggested to play an important role in axonal transport^49,50^. Despite being strongly coexpressed with high confidence ASD genes and having similar characteristics to other ASD genes, it has not been implicated in ASD. Given the infertility phenotypes, this gene may have been missed as an ASD contributor simply due to having too few LoF *de novo* variants and thus insufficient power for association.

The ability to proxy CRISPR perturbation with coexpression enables quick *in silico* analyses to better understand transcriptional consequences of disruptive mutations and functional convergence. However, there are several limitations to this strategy. First, most transcriptional data is derived from bulk tissue. This can obscure relevant coexpression patterns given the cellular heterogeneity and different transcriptional backgrounds among different cell-types^51^. With increasingly larger single-cell datasets, there is a path to overcome this issue in the near future. Second, our primary ASD convergence analysis assumes a single underlying convergent pathway, while multiple pathways likely contribute, especially given the heterogeneity in presentation across individuals diagnosed with ASD. The analytical approach described here can be extended to search for multiple convergent pathways, and as the genotype-phenotype association becomes more granular, we may observe differing degrees of convergence for genes contributing to distinct phenotypic components of ASD. Finally, the use of post-mortem samples cannot fully capture convergence during early development. Transcriptional consequences that may affect neurobiology prenatally cannot be easily captured using postnatal tissue. Future investigations assessing how transcriptional convergence differs across a developmental timespan will prove critical to assess the relevance of using stage-specific biospecimens to answer specific biological questions.

In conclusion, coexpression provides an imperfect but simple proxy for context-specific transcriptional consequences of CRISPR perturbation and enables assessing convergence across many risk genes to provide insight into biology of disease. Most notably, this approach may facilitate the identification of novel risk genes not captured by even the best-powered sequencing studies to date.

## Data Availability

Nearly all data are publicly available and sources are described in the manuscript. The few that are not will be made publicly available but earlier access can be made available on request.

## Acknowledgments

C.L. was supported by a Vanier Graduate Scholarship from the Canadian Institutes of Health Research (CIHR). M.M.O. and J.M.F. were supported by Autism Speaks Foundation Postdoctoral Fellowships (#11815 and #11852, respectively). X.N. was supported by F32MH115614) and K99MH121577. G.A.R. was supported by a CIHR Foundation Scheme grant (#332971). This work was also supported by R01MH123155 (DMR, KJB, MET), R01HD096326 (MET), R01NS093200 (MET), R01MH111776 (DMR), and the Simons Foundation for Autism Research (#573206 to MET).

Data were generated as part of the CommonMind Consortium supported by funding from Takeda Pharmaceuticals Company Limited, F. Hoffman-La Roche Ltd and NIH grants R01MH085542, R01MH093725, P50MH066392, P50MH080405, R01MH097276, RO1-MH-075916, P50M096891, P50MH084053S1, R37MH057881, AG02219, AG05138, MH06692, R01MH110921, R01MH109677, R01MH109897,

U01MH103392, U01MH116442, project ZIC MH002903 and contract HHSN271201300031C through IRP NIMH. Brain tissue for the study was obtained from the following brain bank collections: The Mount Sinai/JJ Peters VA Medical Center NIH Brain and Tissue Repository, the University of Pennsylvania Alzheimer’s Disease Core Center, the University of Pittsburgh Brain Tissue Donation Program, and the NIMH Human Brain Collection Core. CMC Leadership: Panos Roussos, Joseph Buxbaum, Andrew Chess, Schahram Akbarian, Vahram Haroutunian (Icahn School of Medicine at Mount Sinai), Bernie Devlin, David Lewis (University of Pittsburgh), Raquel Gur (University of Pennsylvania), Chang-Gyu Hahn (Thomas Jefferson University), Enrico Domenici (University of Trento), Mette A. Peters, Solveig Sieberts (Sage Bionetworks), Stefano Marenco, Barbara K. Lipska, Francis J. McMahon (NIMH). All ASD gene lists were provided from the ASC-SSC Genomics Consortium.

## Data availability

RNAseq covariate and data is available from the CommonMind Consortium at http://CommonMind.org.

