## Supplementary Figures for "Convergent coexpression of autism associated genes suggests some novel risk genes may not be detectable in large-scale genetic studies"

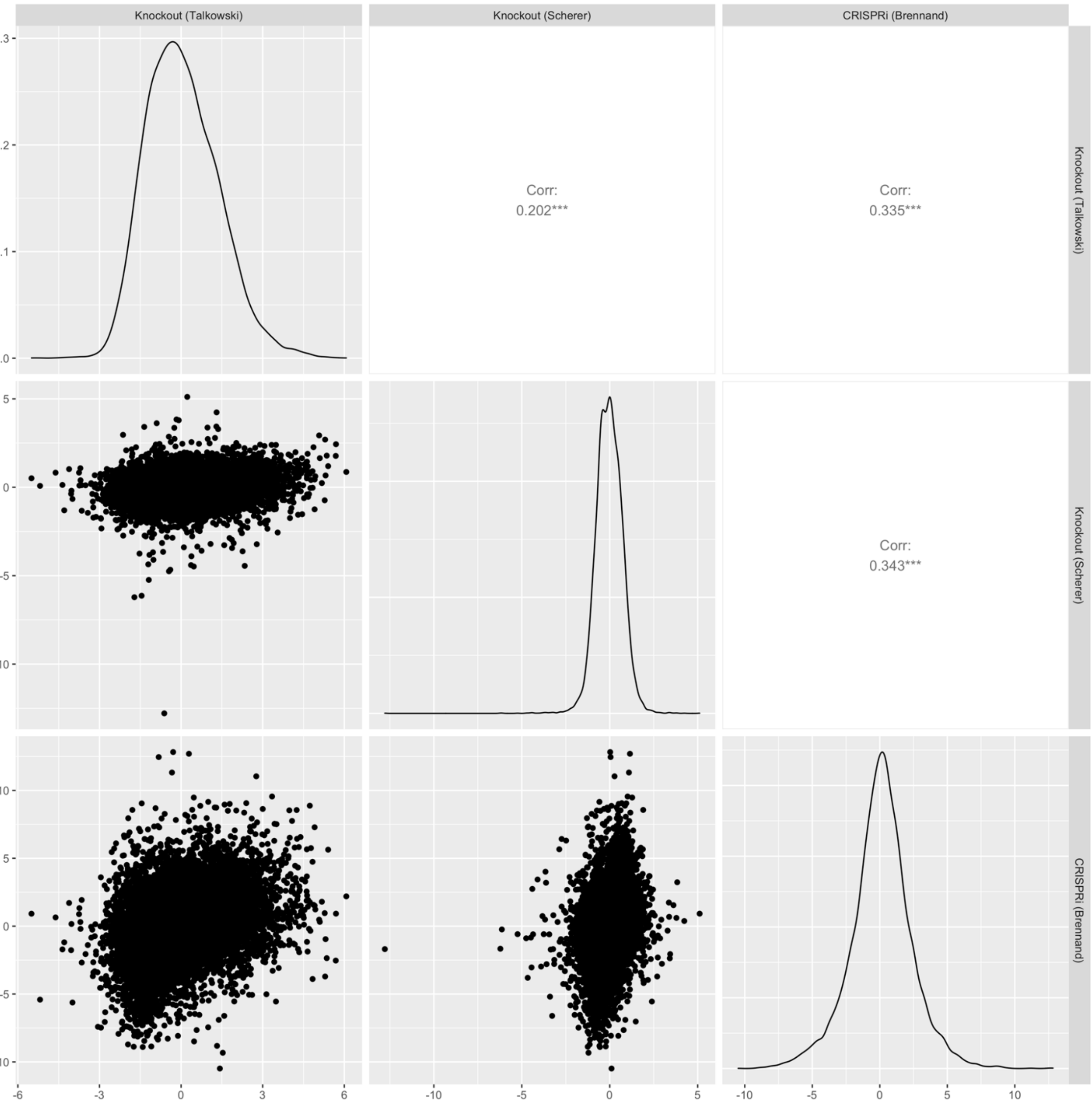


**Supplementary Figure 1. Correlation of differential expression between different *SCN2A* experiments.** Differential expression represented with Z-scores for both axes.

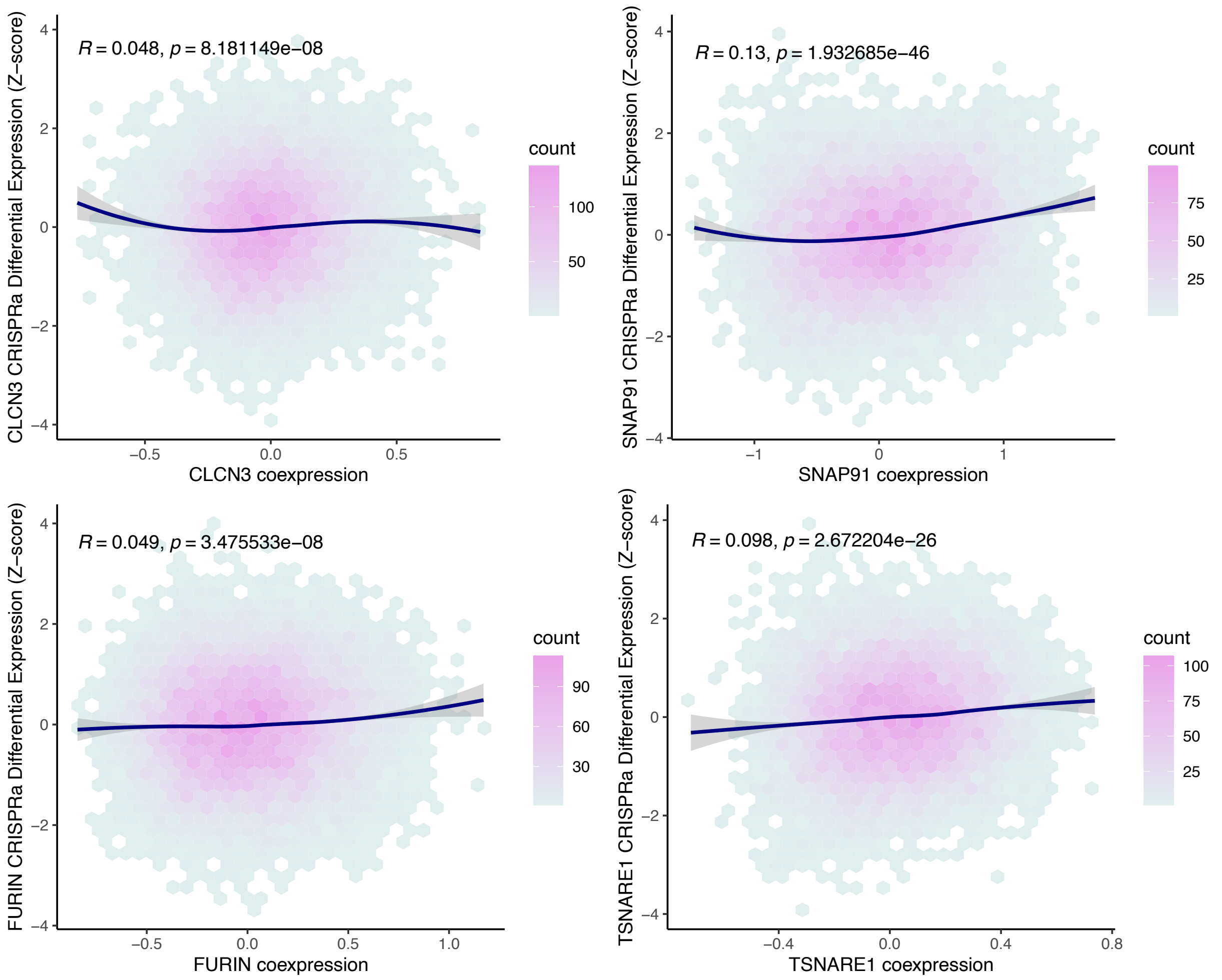


**Supplementary Figure 2. Coexpression positively correlates with differential expression for CRISPR activation.** Coexpression represented as a Fisher transformed Pearson's correlation Z-score. A Pearson correlation was done to assess the correlation between coexpression and differential expression. The curve for each panel was fitted with a locally weighted smoothing (LOESS) regression.

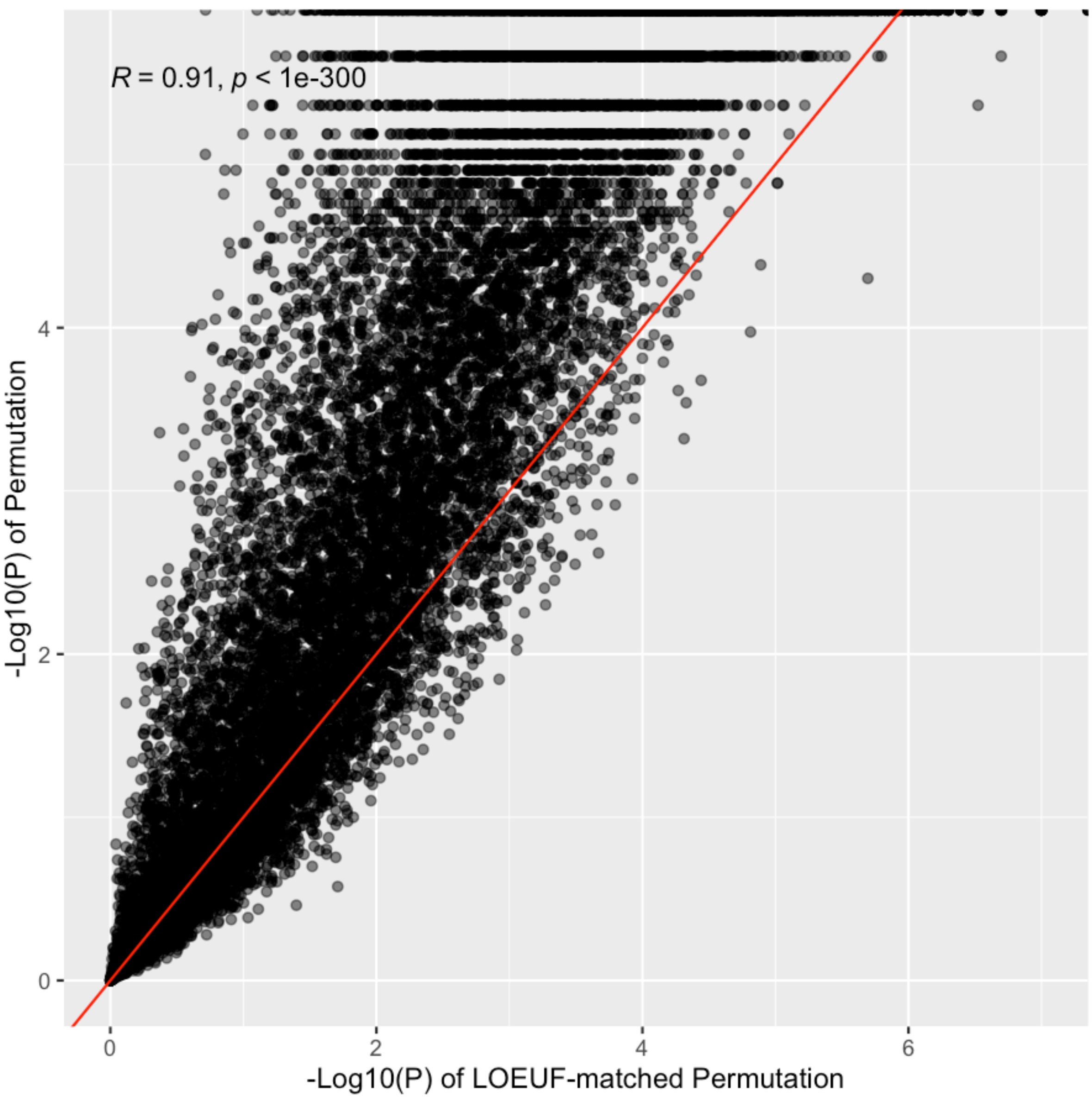


**Supplementary Figure 3. LOEUF-adjusted permutation has similar ranks of genes to unadjusted.** Correlation was done using Spearman's rank correlation.

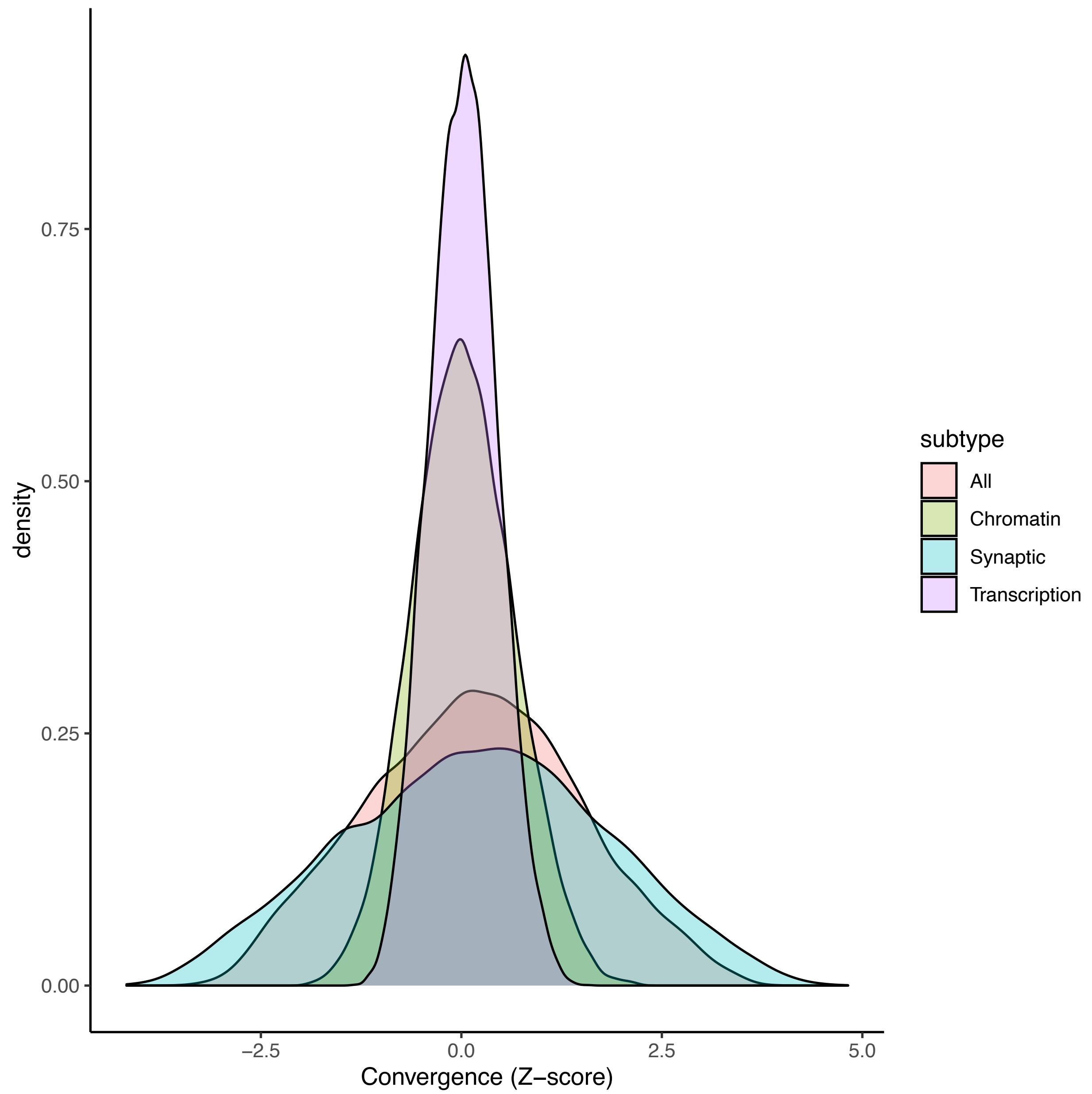


**Supplementary Figure 4. Transcriptional convergence is strongly driven by synaptic genes.**

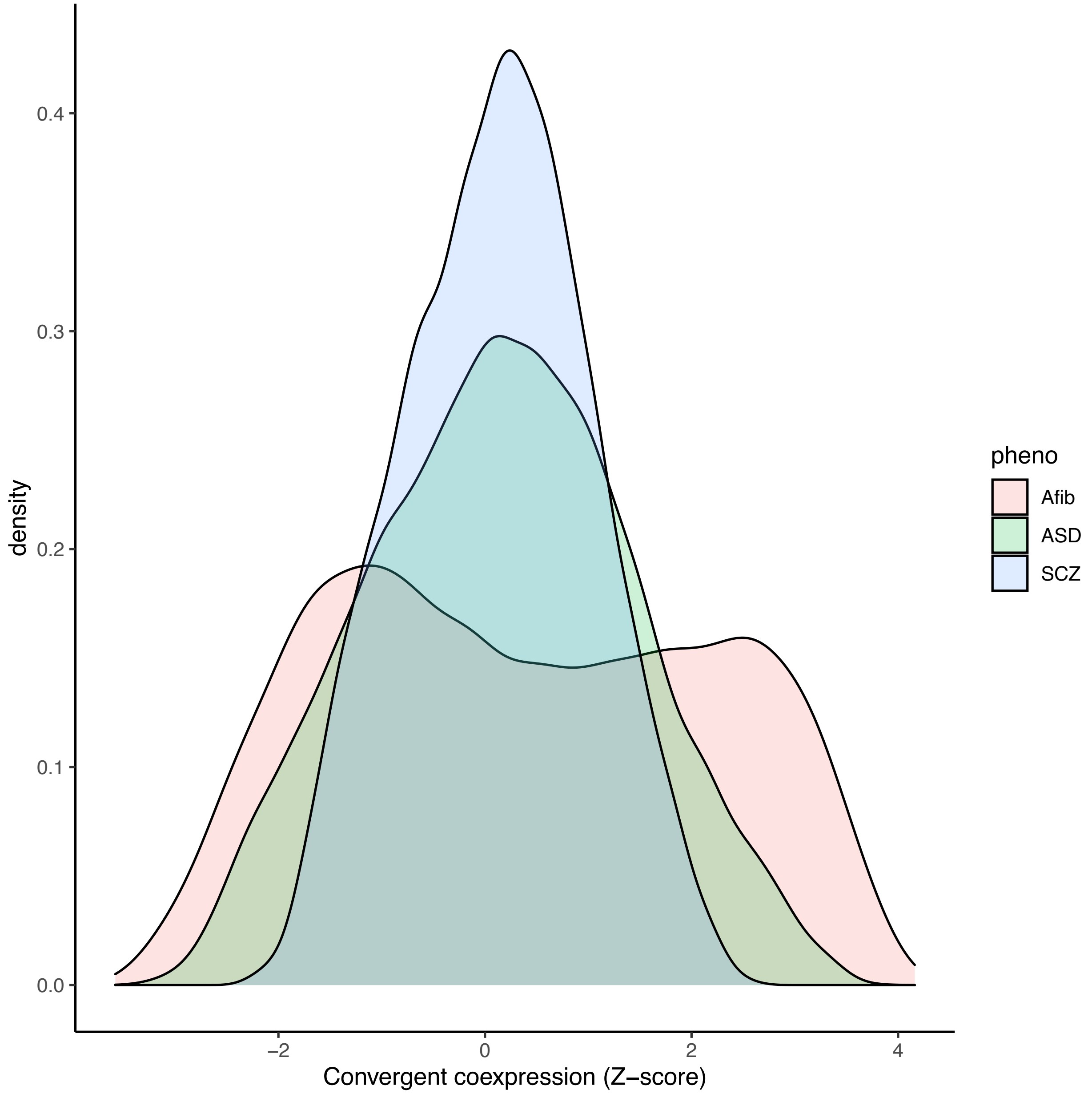


**Supplementary Figure 5. Density distribution of convergent coexpression effect sizes for different traits with respective disease tissue.** For ASD and SCZ, the DLPFC tissue was used. For Afib, the left ventricle was used.

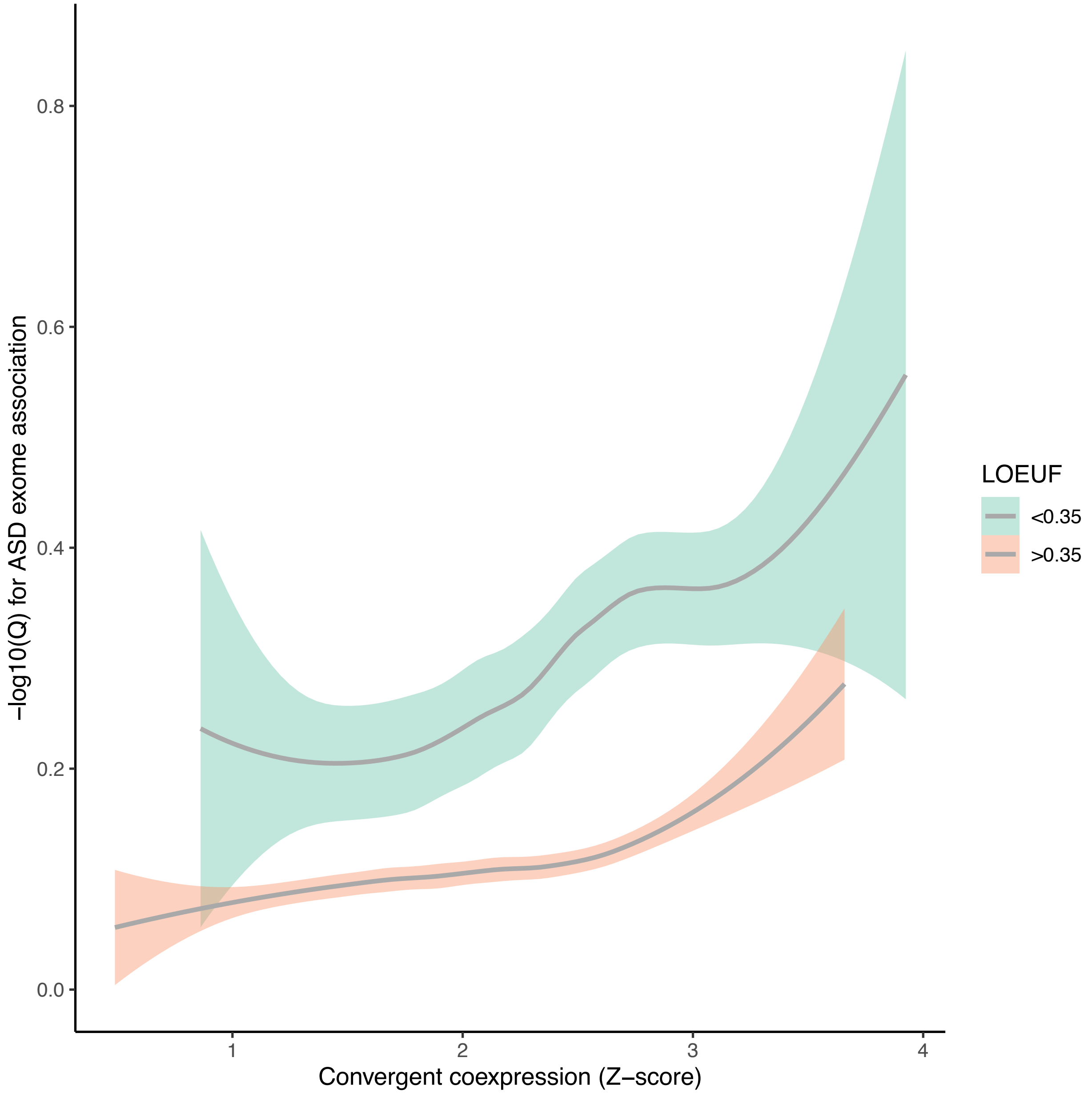


**Supplementary Figure 6. Convergence identifies both tolerant and intolerant genes associated with ASD.**
